# Sex differences in health-related quality of life and poverty risk among older people living with HIV in Spain: a cross-sectional study

**DOI:** 10.1101/2023.11.27.23299048

**Authors:** Néstor Nuño, Alberto Martínez, Susana Martínez, Marta Cobos, Juan Sebastián Hernández, Rosa Polo

## Abstract

**Background:** Current antiretroviral therapies have increased the life expectancy of people living with HIV (PLHIV). There is, however, limited evidence regarding the health-related quality of life (HRQoL) and living conditions of older people living with HIV (OPLHIV) in Spain.

**Methods:** We implemented a self-administered online questionnaire to identify sex differences in HRQoL and poverty risk among Spanish OPLHIV (PLHIV ≥50 years). Participants were contacted through non-governmental organisations. We used the standardised WHOQoL-HIV BREF questionnaire and the Europe 2020 guidelines to estimate HRQoL and poverty risk respectively. The statistical analysis included multivariable generalised linear models with potential confounding variables and robust estimates.

**Results:** The study included 247 OPLHIV (192 men and 55 women). On the WHOQoL-HIV BREF questionnaire, men scored higher on 84% of items and in all six domains. Women had significantly lower HRQoL in five domains: physical health (β: −1.5; 95% CI: −2.5, −0.5; *p*: 0.002), psychological health (β: −1.0; 95% CI: −1.9, −0.1; *p*: 0.036), level of independence (β: −1.1; 95% CI: −1.9, −0.2; *p*: 0.019), environmental health (β: −1.1; 95% CI: −1.8, −0.3; *p*: 0.008), and spirituality/personal beliefs (β: −1.4; 95% CI: −2.5, −0.3; *p*: 0.012). No statistical differences were found in the domain of social relations. Poverty risk was considerable for both men (30%) and women (53%), but women were significantly more likely to experience it (OR: 2.9; 95% CI: 1.3, 6.5; *p*: 0.009).

**Conclusion:** The aging of PLHIV is a public health concern. Our findings indicate that HRQoL and poverty risk among Spanish OPLHIV differ significantly by sex. Spain should, therefore, implement specific policies and interventions to address OPLHIV needs. The strategies must place a high priority on the reduction of sex inequalities in HRQoL and the enhancement of the structural conditions in which OPLHIV live.

## Introduction

Antiretroviral therapy (ART), has transformed the infection of the human immunodeficiency virus (HIV) into a chronic disease, increasing the life expectancy of people living with HIV (PLHIV) at comparable levels to those of the general population [1, 2]. Several studies found relatively positive health-related quality of life (HRQoL) for PLHIV in the ART era [3]. However, levels remained lower than those in the general population [4–6] and people with other chronic diseases [7].

Long-term HIV infection is a critical factor influencing HRQoL of PLHIV, as comorbidities, age-related impediments and non-infectious diseases are greatly prevalent among PLHIV [8. 9]. Nevertheless, multidisciplinary approaches emphasise the importance of addressing HIV-related impacts also from social and structural perspectives [10, 11]. According to various studies, HRQoL of PLHIV is negatively impacted by socio-demographic and psychological factors such as stigma, unwanted loneliness, depression, and financial stress [12–16].

Because of the early aging of the immune system caused by HIV infection, the literature consensus refers PLHIV who are ≥50 years as older people living with HIV (OPLHIV) [17]. As of 2016, there were 5.7 million OPLHIV in the world, and it was estimated a substantial increase in the coming decades [12, 14]. Several studies have confirmed that HRQoL of OPLHIV is lower than younger PLHIV [18–20]. Approximately 53% of PLHIV in Spain are OPLHIV, 15% are classified as frail, and 57% rated their health as poor or very poor in the previous year [21]. It should be noted, however, that none of these estimates take sex differences into account.

PLHIV experiences are closely associated with poverty, which encompasses material (e.g., the access to consumer goods), labour and economic dimensions. PLHIV at working age typically have greater difficulty finding employment due to HIV-related stigma. In turn, their jobs generally have worse conditions and salaries [22–24]. The structural inequalities that PLHIV experience over time result in deprivation, low social support and poor pensions in old age [25–28].

Several studies have described the HRQoL and structural living conditions of OPLHIV in different countries [29, 30]. However, these dimensions remain understudied in Spain, as only one study has assessed the HRQoL of OPLHIV, but without accounting for sex differences [18], and none has examined the structural conditions in which OPLHIV live. This cross-sectional study aims to identify sex differences in HRQoL and poverty risk among Spanish OPLHIV.

## Materials and methods

### Study design

We conducted a cross-sectional study between February and October 2022. A self-administered questionnaire that collected socio-demographic, health, lifestyle, and HRQoL information was implemented online. PLHIV ≥50 years in Spain who were able to make decisions autonomously were invited to participate in the study. Our study complies with the guidelines for strengthening the reporting of observational studies in epidemiology (STROBE). The STROBE is a 22-item checklist developed to facilitate the understanding of the methods, data analysis and results of observational studies [31].

### Sample size and participant selection

We calculated a non-probabilistic minimum sample size of 222 participants. Differences in HIV incidence in Spain were considered to estimate participants by sex. According to the last Spanish epidemiological surveillance report on HIV and AIDS, 84% of HIV cases were reported in men and 16% in women. For decades, HIV cases by sex have remained consistent. Applying a proportionate stratification criterion, the number of participants by sex was 186 men and 36 women. Sample size calculations are described in S1 File. Participants were selected by convenience in collaboration with gTt-VIH, one of the largest non-governmental organisations (NGOs) in Spain working with PLHIV. In order to identify participants who met the inclusion criteria, gTt-VIH collaborated with 19 smaller NGOs throughout the country.

### Outcomes and instruments

The main outcomes were self-reported HRQoL and poverty risk. We assessed HRQoL using the standardised WHOQoL-HIV BREF questionnaire. This instrument was specifically designed for PLHIV [32] and was recently validated in Spain [18]. We selected the WHOQoL-HIV BREF over other HRQoL instruments because of its briefness and robust psychometric properties [33]. In addition, the WHOQoL-HIV BREF has previously been applied to OPLHIV in Spain, France and Portugal [18, 34, 35]. The WHOQoL-HIV BREF questionnaire consists of 31 items grouped into six domains – physical health, psychological health, level of independence, social relations, environmental health, and spirituality/personal beliefs. The questions are scored on a five-point Likert scale between 1 and 5. For each domain, scores range from 4 to 20. Higher scores are associated with better HRQoL. Measurements were performed according to the questionnaire instructions. The Spanish validation of the WHOQoL-HIV BREF questionnaire showed an acceptable internal consistency with a Cronbach’s alpha coefficient of 0.70 and McDonald’s omega coefficient of 0.80 across most domains [18]. In our study, internal consistency was also acceptable, with a global Cronbach’s alpha coefficient of 0.86 and a global McDonald’s omega coefficient of 0.88. The psychometric properties of the WHOQoL-HIV BREF questionnaire by domain are presented in S2 File.

Poverty risk was defined, according to the Europe 2020 strategy guidelines [36], as a person with low labour intensity, low income or severe material deficiencies. People with low work intensity are those with an employment intensity below 20% of their total work potential in the year prior to the questionnaire. Work potential is the quotient between the number of months in which the person worked, and the number of months the person did not work without force majeure impediments. This indicator is not applicable to people >60 years. People with low income are those who received incomes below 60% of the national median equivalent income in the previous year. According to the Spanish National Institute of Statistics, the equivalent median income in 2020 was €16,339 for men and €15,804 for women [37]. People with severe material deprivation lack at least 4 (out of 9) dimensions described in the Europe 2020 strategy [38].

### Data collection and analysis

Data was collected using the open-source app Kobo Toolbox (KoBo, Inc.). To reduce missing data, all questions were mandatory, but participants were permitted to decline to answer or select a neutral response. In order to prevent the same user from submitting the questionnaire twice, we programmed the tool to accept only one response per electronic device. Upon completing data collection, we reviewed and cleaned the data for errors and inconsistencies in the responses. We used descriptive statistics (e.g., frequency tables for categorical variables, and mean and standard deviation for numerical variables) to present socio-demographic, health, and lifestyle information. To identify differences across numerical variables by sex, we used the Student’s t-test or its non-parametric alternative, the Mann-Whitney test. In the case of categorical variables, we applied the chi-square test or Fischer’s exact test.

To assess sex differences in HRQoL domains and poverty risk, we used univariable generalised linear models with sex (0=men; 1=women) as a predictor variable. Additionally, we designed multivariable generalised linear models incorporating potential confounding variables derived from the literature. Multivariable models for HRQoL domains included the following predictor variables: sex, age, years since HIV diagnosis, being on ART (0=no; 1=yes), number of comorbidities experienced in the previous year or chronic^1^ and presence of self-reported mobility impairments (0=no; 1=yes) and mental disorders (0=no; 1=yes) experienced in the previous year or chronic. The multivariable model for poverty risk included the following predictor variables: sex, region of origin (0=other; 1=Spain), years since HIV diagnosis, employment status (0=other; 1=employed), education level (0=mandatory; 1=higher), relationship status (0=single; 1=stable partner), number of comorbidities experienced in the previous year or chronic, and presence of self-reported mobility impairments and mental disorders experienced in the previous year or chronic. We used robust estimates of standard errors to calculate confidence intervals. The statistical analysis was performed using R 3.6 (R Project for statistical computing).

### Ethics

Participation in the study was voluntary and no financial compensation was offered to encourage participation. The researchers did not receive any compensation for their work on the study. Due to the observational and non-invasive nature of the study, participation did not entail any risk.

Neither the questionnaires nor informed consent forms collected any personal information as some PLHIV in Spain refuse to disclose their serological status for HIV publicly due to HIV-related stigma [39]. The ethical board of La Princesa Hospital (Madrid) exempted the study from review. The study was conducted in accordance with the Helsinki Declaration. All participants signed an informed consent form prior to completing the questionnaire.

## Results

### Participant characteristics

The questionnaire was completed by 257 participants, but 10 participants were excluded from the final analysis: n=3 did not indicate their age and n=7 were ≤50 years. The final analysis included 247 participants: 192 men (78%) and 55 women (22%). Seventy-three percent of male (n=140) and 85% of female (n=47) participants were ≤60 years.

In the study, 89% of Spanish regions were represented, with the majority of participants hailing from Madrid (23%), Catalonia (22%), Valencia (9%), and Andalusia (9%) regions. Five participants (2%) did not indicate their residence region. Fig 1 represents the distribution of participants by region.

**Figure 1.**
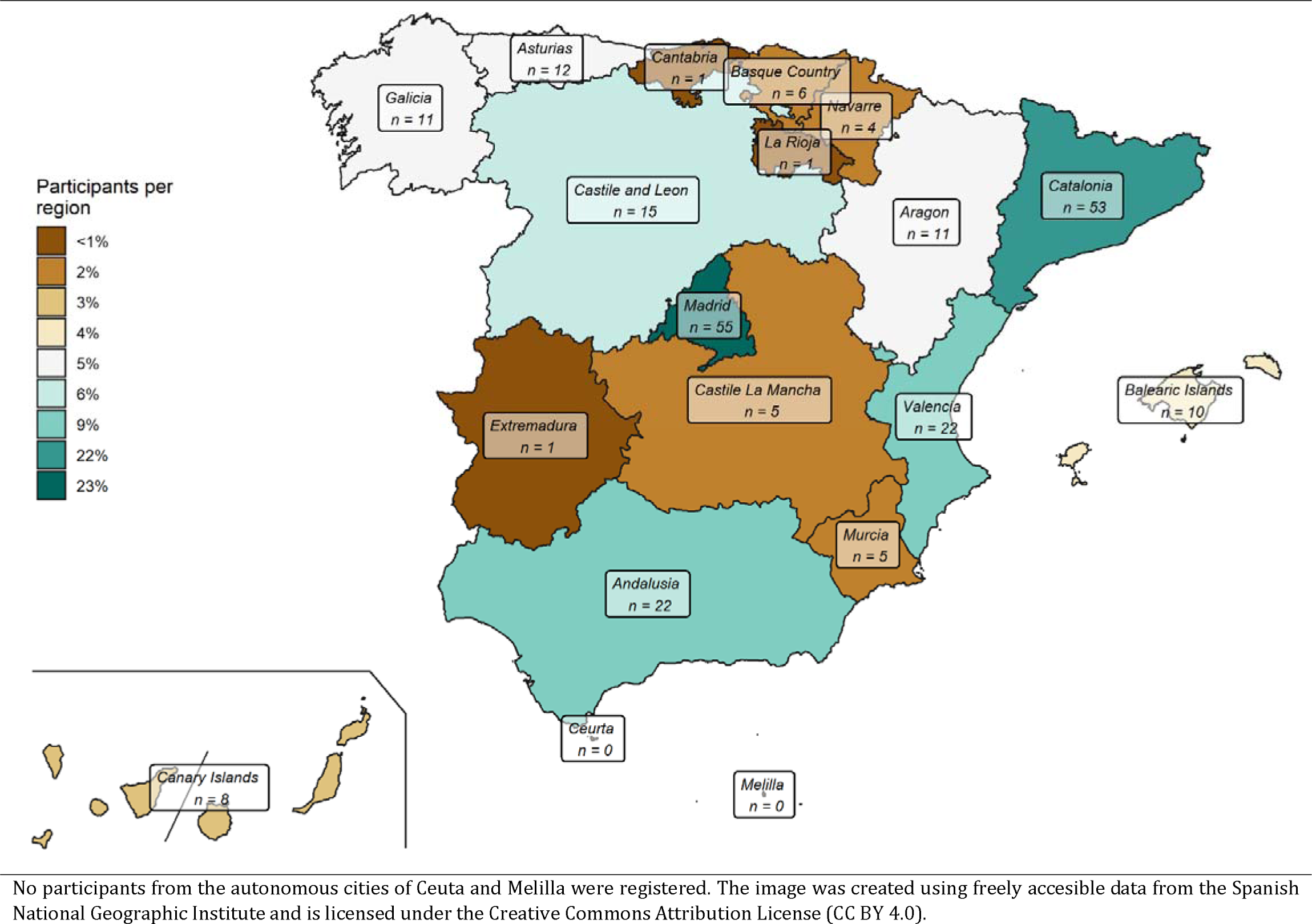
Distribution of study participants by region

Table 1 summarises the sociodemographic and health characteristics of participants. The mean age was 56.9 (SD=5.1) years, the majority (89%) was born in Spain, and 83% held a higher education degree, with a mean of 14.5 (SD=3.9) years of study. Fifty-seven percent of participants identified themselves as homosexuals, 41% had a stable partner and 56% were employed. Concerning health characteristics, 56% of participants self-reported being in good health in the previous year. The average number of years since HIV diagnosis was 21.2 (SD=10.5). Almost all participants had an undetectable viral load (97%) and were on ART (99%). Seventy-four percent of HIV infections result from unprotected sex. Mobility impairments (21%), mental disorders (20%) and renal failure (5%) were the most common self-reported comorbidities experienced in the previous year or chronic. An average of 1.2 (SD=1.1) comorbidities were reported by participants, 35% smoked, 79% consumed alcohol, 82% exercised regularly, and 41% had used some drugs^2^ during the previous month.

**Table 1.** Socio-demographic, health and lifestyle characteristics of study participants by sex.

|  | Total |  | Men |  | Women |  | p-value |
| --- | --- | --- | --- | --- | --- | --- | --- |
|  | N | % (n) / Mean (SD) | N | % (n) / Mean (SD) | N | % (n) / Mean (SD) |  |
| *Socio-demographic characteristics* | 247 |  | 192 |  | 55 |  |  |
| Age (years) |  | 56.9 (5.1) |  | 57.1 (5.4) |  | 56.3 (4.0) | 0.606 |
| Birthplace (Spain) |  | 88.7% (219) |  | 86.5% (166) |  | 96.4% (53) | 0.041 |
| Degree in education (higher education)<sup>a</sup> |  | 82.6% (204) |  | 88.3% (165) |  | 64.5% (36) | <0.001 |
| Education (number of years of study)<sup>b</sup> |  | 14.5 (3.9) |  | 15.1 (3.6) |  | 12.4 (4.2) | <0.001 |
| Sexual orientation (homosexual) | 244 | 56.6% (138) | 190 | 70.5% (134) | 54 | 7.4% (4) | <0.001 |
| Civil state (stable partner) | 246 | 41.1% (101) | 191 | 43.5% (83) |  | 32.7% (18) | 0.154 |
| Working situation (salaried employee) | 245 | 56.3% (138) | 190 | 59.0% (112) |  | 47.3% (26) | 0.124 |
| *Health characteristics* | 247 |  | 192 |  | 55 |  |  |
| Self-reported good health (previous year) |  | 55.6% (138) |  | 62.0% (119) |  | 34.6% (19) | <0.001 |
| Years since HIV diagnosis | 236 | 21.2 (10.5) | 181 | 19.2 (10.0) |  | 28.0 (9.4) | <0.001 |
| Viral load (undetectable) | 243 | 97.1% (236) | 189 | 97.9% (185) | 54 | 94.4% (51) | 0.183 |
| ART | 246 | 99.2% (244) | 191 | 99.5% (190) |  | 98.2% (54) | 0.346 |
| HIV infection route (unprotected sex) | 245 | 73.5% (180) | 190 | 80.0% (152) |  | 50.9% (28) | <0.001 |
| Mobility impairments<sup>c</sup> | 245 | 21.2% (52) | 191 | 17.3% (33) | 54 | 35.2% (19) | 0.004 |
| Mental disorders<sup>c</sup> | 245 | 19.6% (48) | 191 | 19.4% (37) | 54 | 20.4% (11) | 0.870 |
| Renal failure<sup>c</sup> | 245 | 4.5% (11) | 191 | 2.1% (4) | 54 | 13.0% (7) | 0.001 |
| Comorbidities (total number)<sup>c</sup> | 245 | 1.2 (1.1) | 191 | 1.2 (1.1) | 54 | 1.4 (1.2) | 0.176 |
| *Lifestyle characteristics* | 247 |  | 192 |  | 55 |  |  |
| Smoke | 246 | 35.4% (87) | 191 | 34.0% (65) |  | 40.0% (22) | 0.415 |
| Alcohol consumption | 245 | 79.2% (194) | 190 | 81.6% (155) |  | 70.9% (39) | 0.086 |
| Regular physical activity |  | 82.2% (203) |  | 83.9% (161) |  | 76.4% (42) | 0.201 |
| Drugs consumption (previous month) |  | 41.3% (102) |  | 44.8% (86) |  | 29.1% (16) | 0.037 |

SD: standard deviation. ART: antiretroviral treatment. p-values calculated applying the chi-square, Fisher's exact and Mann-Whitney tests.
<sup>a</sup>Higher education in Spain is not compulsory and may require a fee.
<sup>b</sup>It includes the following categories: primary education (6 years); secondary education (10 years), baccalaureate (12 years); higher education, non-university (14 years); higher education, university (17 years); and higher education, master/doctorate (20 years).
<sup>c</sup>Self-reported in the previous year or chronic.

As for socio-demographic characteristics, women had significantly lower levels of education (*p*<0.001), fewer years of study (*p*<0.001), and identified themselves as homosexuals less often (*p*<0.001). In contrast, women were born primarily in Spain (*p*=0.041). Across health and lifestyle characteristics, women had significantly lower perceptions of good health (*p*<0.001), higher years since HIV diagnosis (*p*<0.001), lower percentages of infection through unprotected sex (*p*<0.001), greater self-reported mobility impairments (*p*=0.004) and renal failure (*p*=0.001), and less drugs consumed (*p*=0.037) (Table 1).

### Health-related quality of life

In the WHOQoL-HIV BREF questionnaire, men scored higher than women on 26 of the 31 items (84% of the total). In two items (overall general health, and information for daily living) men and women scored similarly. Women outperformed men on three items: social support, personal relationships, and death and dying. Men obtained higher scores in the six domains of the WHOQoL-HIV BREF questionnaire (difference range: 2–14%) (Table 2).

**Table 2.** Results of the WHOQoL-HIV BREF questionnaire by sex.

|  | Total (N=247) | Men (N=192) | Women (N=55) |
| --- | --- | --- | --- |
|  | Mean (SD) | Mean (SD) | Mean (SD) |
| *Overall HRQoL/General Health<sup>a</sup>* | 12.6 (2.7) | 12.8 (2.6) | 11.9 (3.1) |
| Overall HRQoL | 3.4 (0.9) | 3.5 (0.9) | 3.0 (0.9) |
| Overall general health | 2.9 (1.0) | 2.9 (1.0) | 2.9 (1.0) |
| *Physical health* | 13.0 (3.4) | 13.4 (3.3) | 11.5 (3.4) |
| Pain and discomfort<sup>b</sup> | 3.5 (1.2) | 3.6 (1.2) | 3.1 (1.2) |
| Symptoms of HIV<sup>b</sup> | 3.8 (1.2) | 3.9 (1.2) | 3.3 (1.2) |
| Energy and fatigue | 3.1 (1.0) | 3.2 (0.9) | 2.9 (1.0) |
| Sleep and rest | 2.6 (1.1) | 2.7 (1.1) | 2.2 (1.0) |
| *Psychological health* | 12.5 (3.1) | 12.7 (3.1) | 11.8 (2.8) |
| Positive feelings | 3.3 (1.0) | 3.3 (1.0) | 3.2 (1.0) |
| Concentration ability | 3.0 (1.0) | 3.0 (0.9) | 2.8 (1.1) |
| Body image self-acceptance | 3.3 (1.0) | 3.4 (1.0) | 3.0 (1.1) |
| Self-satisfaction | 3.2 (1.0) | 3.3 (1.0) | 3.1 (1.0) |
| Negative feelings<sup>b</sup> | 2.9 (1.0) | 2.9 (1.0) | 2.7 (0.9) |
| *Level of independence* | 13.2 (3.2) | 13.5 (3.1) | 12.2 (3.2) |
| Dependence on medication<sup>b</sup> | 3.0 (1.3) | 3.1 (1.3) | 2.8 (1.2) |
| Mobility | 3.2 (1.0) | 3.3 (1.0) | 2.9 (1.0) |
| Activities of daily living | 3.1 (1.0) | 3.2 (1.0) | 2.8 (1.1) |
| Work capacity | 3.9 (1.0) | 3.9 (0.9) | 3.8 (1.1) |
| *Social relations* | 12.5 (3.2) | 12.5 (3.3) | 12.3 (2.9) |
| Social support | 3.3 (1.1) | 3.2 (1.1) | 3.6 (1.0) |
| Sexual satisfaction | 2.4 (1.1) | 2.5 (1.1) | 2.0 (1.1) |
| Personal relationships | 3.1 (1.0) | 3.1 (1.0) | 3.2 (1.0) |
| Social inclusion | 3.7 (0.9) | 3.7 (0.9) | 3.5 (0.9) |
| *Environmental health* | 13.9 (2.6) | 14.2 (2.6) | 13.2 (2.5) |
| Physical safety and security | 3.1 (0.9) | 3.2 (0.9) | 3.0 (0.8) |
| Physical environment | 3.5 (0.8) | 3.5 (0.8) | 3.4 (0.9) |
| Financial resources | 3.3 (1.0) | 3.4 (1.0) | 2.8 (0.9) |
| Information for daily living | 3.7 (0.9) | 3.7 (0.9) | 3.7 (0.8) |
| Participation in leisure activities | 3.4 (1.0) | 3.5 (1.0) | 3.2 (1.0) |
| Home environment | 3.7 (1.0) | 3.7 (1.0) | 3.6 (1.1) |
| Accessibility of health services | 3.5 (1.0) | 3.6 (1.0) | 3.3 (1.1) |
| Transport | 3.6 (1.0) | 3.6 (0.9) | 3.3 (1.1) |
| *Spirituality/personal beliefs* | 12.9 (3.7) | 13.2 (3.8) | 12.1 (3.1) |
| Personal life meaning | 3.4 (1.1) | 3.4 (1.2) | 3.2 (1.0) |
| Forgiveness and blame<sup>b</sup> | 3.1 (1.6) | 3.3 (1.6) | 2.6 (1.5) |
| Concerns about the future<sup>b</sup> | 3.0 (1.2) | 3.0 (1.2) | 2.8 (1.1) |
| Death and dying<sup>b</sup> | 3.5 (1.2) | 3.4 (1.2) | 3.5 (1.1) |

SD: standard deviation.
<sup>a</sup>According to the instructions for the questionnaire, these items do not belong to any domain.
<sup>b</sup>Reversed items recorded.

### Poverty risk

A major material deprivation among participants was the inability to pay for unexpected expenses (37%), the capacity to afford a car (29%), and the inability to take an annual week off (23%). The average number of months participants were unemployed was 3.9 (SD=5.0), compared to 7.6 (SD=5.0) months they worked. Approximately 33% of respondents experienced difficulties making ends meet, with the median income reported at €20,000 (IQR=€14,000, €34,000). In total, 13% of participants suffered severe material deprivation, 38% had low work intensity, and 21% earned a low income. The percentage of participants at risk of poverty was 35% (Table 3).

**Table 3.** Material, labour and economic deprivation and poverty risk by sex.

|  | Total |  | Men |  | Women |  | *p*-value |
| --- | --- | --- | --- | --- | --- | --- | --- |
|  | N | % (n) /<br>Mean (SD) /<br>Median<br>(IQR) | N | % (n) /<br>Mean (SD) /<br>Median<br>(IQR) | N | % (n) /<br>Mean (SD) /<br>Median<br>(IQR) |  |
| *Material deprivation<sup>a</sup>* | 247 |  | 192 |  | 55 |  |  |
| Consume meat or fish (each two days) | 244 | 5.7% (14) | 189 | 5.8% (11) |  | 5.5% (3) | 0.918 |
| Car | 235 | 28.9% (68) | 181 | 24.3% (44) | 54 | 44.4% (24) | 0.004 |
| Maintain an adequate household temperature | 243 | 15.2% (37) | 188 | 11.2% (21) |  | 29.1% (16) | 0.001 |
| Pay for unexpected expenses<sup>b</sup> | 239 | 36.8% (88) | 186 | 30.1% (56) | 53 | 60.4% (32) | <0.001 |
| Washing machine | 245 | 2.0% (5) | 190 | 2.1% (4) |  | 1.8% (1) | 1.000 |
| At least one payment delay (previous year) | 242 | 14.1% (34) | 188 | 13.3% (25) | 54 | 16.7% (9) | 0.530 |
| Telephone | 246 | 0.8% (2) | 191 | 1.0% (2) |  | 0.0% (0) | 1.000 |
| Television | 246 | 4.1% (10) | 191 | 3.7% (7) |  | 5.5% (3) | 0.697 |
| Have a week of holidays (per year) | 174 | 23.0% (40) | 138 | 18.8% (26) | 36 | 38.9% (14) | 0.011 |
| *Labour deprivation<sup>c</sup>* | 187 |  | 140 |  | 47 |  |  |
| Unemployment (months previous year)<sup>d</sup> | 167 | 3.9 (5.0) | 127 | 3.2 (4.7) | 40 | 6.0 (5.3) | 0.007 |
| Employment (months previous year) | 167 | 7.6 (5.0) | 127 | 8.2 (4.8) | 40 | 5.7 (5.3) | 0.019 |
| *Economic deprivation* | 247 |  | 192 |  | 55 |  |  |
| Total income (previous year)<sup>e</sup> | 161 | 20 (14, 34) | 130 | 23 (14, 37) | 31 | 17 (8, 20) | 0.001 |
| Make ends meet (with any difficulty) |  | 33.2% (82) |  | 27.6% (53) |  | 52.7% (29) | <0.001 |
| *Poverty risk estimation* | 247 |  | 192 |  | 55 |  |  |
| Severe material deprivation |  | 12.6% (31) |  | 10.9% (21) |  | 18.2% (10) | 0.153 |
| Low work intensity<sup>b</sup> | 167 | 38.3% (64) | 127 | 31.5% (40) | 40 | 60.0% (24) | 0.001 |
| Low income | 187 | 20.9% (39) | 151 | 17.2% (26) | 36 | 36.1% (13) | 0.012 |
| Poverty risk |  | 35.2% (87) |  | 30.2% (58) |  | 52.7% (29) | 0.002 |

SD: standard deviation; IQR: interquartile range. *p*-values calculated applying the chi-square, Fisher's exact and Mann-Whitney tests.
<sup>a</sup>Difficulties to achieve/afford/have.
<sup>b</sup>Of at least €750.
<sup>c</sup>Participants >60 years are excluded.
<sup>d</sup>Without force majeure impediments.
<sup>e</sup>In thousands of euros.

In general, women reported more material, labour, and economic deprivation than men. We found statistically differences in the capacity to afford a car (*p*=0.004), keeping an adequate household temperature (*p*=0.001), paying unexpected expenses (*p*<0.001), and having an annual week of holidays (*p*=0.011). Similarly, women reported fewer months working (*p*=0.019) and less income (*p*=0.001), as well as a higher percentage of unemployed months (*p*=0.007) and difficulty making ends meet (*p*<0.001). The prevalence of low working intensity (*p*=0.001), low income (*p*=0.012) and poverty risk (*p*=0.002) was also higher among women (Table 3).

### Sex differences in health-related quality of life and poverty risk

The multivariable statistical analysis showed that women had significantly lower HRQoL in five domains: physical health (β: −1.5; 95% CI: −2.5, −0.5; *p*: 0.002), psychological health (β: −1.0; 95% CI: −1.9, −0.1; *p*: 0.036), level of independence (β: −1.1; 95% CI: −1.9, −0.2; *p*: 0.019), environmental health (β: −1.1; 95% CI: −1.8, −0.3; *p*: 0.008), and spirituality/personal beliefs (β: −1.4; 95% CI: −2.5, −0.3; *p*: 0.012). No statistical differences were found in the domain of social relations. Women were 2.9 times more likely to experience poverty risk than men (OR: 2.9; 95% CI: 1.3, 6.5; *p*: 0.009) (Table 4).

**Table 4.** Statistical models for assessing sex differences in HRQoL and poverty risk.

|  | Unadjusted model<sup>a</sup> |  |  | Adjusted model<sup>b</sup> |  |  |
| --- | --- | --- | --- | --- | --- | --- |
|  | Mean difference | 95% CI | p-value | Mean difference | 95% CI | p-value |
| Physical health | -1.9 | -2.9, -0.9 | <0.001 | -1.5 | -2.5, -0.5 | 0.002 |
| Psychological health | -0.9 | -1.8, -3e<sup>-2</sup> | 0.027 | -1.0 | -1.9, -0.1 | 0.036 |
| Level of independence | -1.3 | -2.2, -0.3 | 0.008 | -1.1 | -1.9, -0.2 | 0.019 |
| Social relations | -0.2 | -1.1, 0.6 | 0.585 | -0.4 | -1.3, 0.5 | 0.378 |
| Environmental health | -1.0 | -1.8, -0.2 | 0.010 | -1.1 | -1.8, -0.3 | 0.008 |
| Spirituality/personal beliefs | -1.1 | -2.1, -0.1 | 0.024 | -1.4 | -2.5, -0.3 | 0.012 |
|  | Unadjusted model<sup>a</sup> |  |  | Adjusted model<sup>c</sup> |  |  |
|  | OR | 95% CI | p-value | OR | 95% CI | p-value |
| Poverty risk | 2.6 | 1.4, 4.8 | 0.003 | 2.9 | 1.3, 6.5 | 0.009 |

CI: confidence interval; OR: odd ratio.
<sup>a</sup>Estimated by generalised linear models with robust estimates.
<sup>b</sup>Estimated by generalised linear models with robust estimates adjusted by age, years since HIV diagnosis, being on ART, number of comorbidities and presence of self-reported mobility impairments and mental health disorders.
<sup>c</sup>Estimated by generalised linear models with robust estimates adjusted by region of origin, years since HIV diagnosis, employment status, education level, relationship status, number of comorbidities and presence of self-reported mobility impairments and mental health disorders.

## Discussion

We conducted a cross-sectional study with 247 Spanish older people living with HIV (OPLHIV) to identify sex differences in their health-related quality of life (HRQoL) and poverty risk. HRQoL was determined using the standardised WHOQoL-HIV BREF questionnaire, which was recently validated in Spain [18]. Poverty risk was estimated based on the guidelines of the Europe 2020 strategy, which is the current methodology used in the European Union. This is, to our knowledge, one of the few studies that assess HRQoL and poverty risk for OPLHIV in Spain using standardised instruments and accounting for sex differences.

Our results showed that male and female participants differed in terms of socio-demographic, health, and structural characteristics. In general, women defined themselves as homosexuals less often, and had lower levels of education (degrees completed and years of study), income, work intensity, drug consumption, and self-reported perception of good health in the previous year. Conversely, women had greater self-reported mobility impairments and renal failure, more years since HIV diagnosis, higher levels of material deprivation, and more difficulties to make ends meet. These characteristics are consistent with those of previous studies with OPLHIV [40–42].

We found substantial differences in HRQoL by sex. Women had significantly lower HRQoL in the domains of physical health, psychological health, level of independence, environmental health, and spirituality/personal beliefs. Recent studies have reported poor physical health among women living with HIV (WLHIV). In England, Brañas et al., found that WLHIV ≥50 years had a lower HRQoL than their male counterpart [42]. However, the study was limited to 100 participants (27 women). Additionally, in a cohort of 1,000 OPLHIV (25% women) in Italy it was found that WLHIV had lower physical strength, increased frailty and lower HRQoL than men living with HIV (MLHIV) [43]. Sex differences in psychological health are common among WLHIV, as they generally suffer from depression, anxiety and post-traumatic stress [44]. A study with 357 WLHIV in Ethiopia found that anxiety and depression were present in around 30% of the participants. Factors such as low education, divorce, unemployment or financial burdens were negatively associated with depression. However, the study did not focus on OPLHIV exclusively [45]. In a recent review, Waldron et al., showed that WLHIV generally have strong histories of physical and sexual abuse, caregiving stress, elevated internalised stigma, and a wide range of barriers to care. As a result, there is a high prevalence of depression, anxiety, and trauma-related mental health issues among WLHIV. For example, WLHIV could be up to four times more likely to be diagnosed with major depressive disorder and experience more severe depressive symptom than HIV-seronegative women [46]. However, the review did not focus on sex differences, nor specifically on OPLHIV. The observed living and structural conditions of women in our study may be related to sex differences in the level of independence and environmental health domains, as these two domains include items relating to work capacity, physical environment, living conditions, and economic resources. Other studies also indicate that WLHIV generally live in poorer conditions and have lower socioeconomic status than MLHIV. The study conducted by Kalichman et al., found that income inequality, internalised stigma, and enacted stigma were significantly associated with HIV suppression among WLHIV (but not in MLHIV). However, the study did not focus on OPLHIV exclusively [47]. Likewise, a qualitative study among WLHIV (89% African American) found that that poverty, unemployment, limited access to healthcare resources and stigma impacted negatively on their health and ability to engage in HIV care [48]. This study, however, did not focus on sex differences, nor specifically on OPLHIV.

Our results are also consistent with recent studies in Spain. In a study conducted by Fumaz et al., WLHIV had significantly lower physical function and psychological health than MLHIV, although the sample did not include OPLHIV exclusively [49]. Fuster-Ruiz de Apodaca et al. found that WLHIV scored lower in 61% of items and the six domains of the WHOQoL-HIV BREF questionnaire. Although 38% of participants (n=549) were OPLHIV, sex differences in HRQoL were only examined for the total sample [18]. Ruiz-Algueró et al. found that 63% of OPLHIV described their health as good in the previous year, although the results were not disaggregated by sex [50]. In our study, we found a similar result with 57% of participants defining their health as good in the previous year. Finally, other studies conducted with OPLHIV in Germany, Brazil, the United States, Italy and Portugal concluded that WLHIV tend to have a worse HRQoL than MLHIV [19, 41–43, 51].

In our study, we found that 53% of women were at risk of poverty and that they were more likely to suffer it than men. Despite this difference, it is important to remark that men were also at high poverty risk (30%). Other studies have shown that OPLHIV are particularly vulnerable to poverty and inequality, although none of them specifically address sex differences. In Canada, Sok et al. reported that 87% of 496 PLHIV did not have access to basic needs (e.g., clothing, food). Particularly among OPLHIV, unmet basic needs were associated with poorer physical and psychological health [30]. Similarly, Hessol et al. found that 32% of 230 OPLHIV in the United States lacked regular access to healthy food. In addition, food insecurity was associated with alcohol consumption, sedentary lifestyles, and depression [29]. Finally, a study conducted in the United Kingdom with 307 OPLHIV found that 58% were living at or below the poverty line, only 45% (of those aged 50-64 years) were economically active, and 32% were dependent on social benefits [52].

We identified some limitations in our study. Although our estimates of poverty risk were consistent with previous studies, the impact of the SARS-CoV-2 pandemic on labour and economics could have caused an overestimation of the outcome. For example, the pandemic caused a substantial negative impact on the Spanish economy, with unemployment rates increasing to 36% in April and May 2020 [53]. In contrast, we also observed response patterns that were associated with a potential underestimation of our poverty risk estimates, as questions related to economic, employment, and material deficiencies had a higher proportion of missing responses. There is evidence that socioeconomically disadvantaged groups are more likely to leave these questions unanswered [54, 55] Nevertheless, we do not believe that this missingness alters the interpretation of our results since the proportion of missing responses was similar across sexes. Last but not least, we are unable to generalise our results to the population of OPLHIV in Spain due to the sample size and the selection of participants. Our findings, however, are consistent with those of other previous studies conducted with OPLHIV in Spain.

Overall, our results can be helpful in identifying OPLHIV needs in Spain. According to a recent series published in The Lancet Healthy Longevity, new research areas on HIV and aging are needed to address the complex challenges that OPLHIV face [56]. Commonly, older adults have been marginalised and underrepresented in research [57, 58]. Now it is time to resolve this trend and address how modern societies can meet the needs of elderly populations. There are several factors related to HIV and aging that must be taken into account. In this regard, it will be difficult to achieve tangible and transformative improvements for OPLHIV without gender-sensitive policies and interventions.

## Conclusion

We conducted a cross-sectional study among 247 Spanish older persons living with HIV (192 women and 55 men) in order to identify sex differences in health-related quality of life and poverty risk. In general, women had a significantly lower health-related quality of life and a higher poverty risk than men. In light of these results, future gender-sensitive policies are required to improve living conditions and provide comprehensive care for older people living with HIV in Spain.

## Supporting information

Sample size calculation

Psychometric properties of the WHOQoL-HIV BREF questionnaire

## Data Availability

All data produced in the present study are available upon reasonable request to the authors

## Acknowledgments

We would like to thank all participants for their contribution in the questionnaire. In addition, we also acknowledge gTt-VIH for their assistance in the implementation of the study.

## Author contributions

**Conceptualization:** NN, JSH, RP.

**Data curation:** NN.

**Formal analysis:** NN.

**Funding acquisition:** JSH.

**Investigation:** NN, JSH.

**Methodology:** NN, RP.

**Project administration:** JSH.

**Software:** NN, MC.

**Supervision:** JSH.

**Validation:** AM, SM, RP.

**Visualization:** NN, SM.

**Writing – original draft:** NN.

**Writing – review & editing:** NN, AM, SM, MC, JSH, RP.

## Supporting information

**S1 File.** Sample size calculation.

**S2 File.** Psychometric properties of the WHOQoL-HIV BREF questionnaire.

## Footnotes

1 The full list of comorbidities included: Cardiovascular disease, stroke, diabetes mellitus, respiratory diseases (e.g., pulmonary embolism, coronary angioplasty), hypertension, cancer/tumor, sexually transmitted infection, and dementia/alzheimer.

2 The full lust of drugs included: erection enhancing drugs, cannabis, hashish, cocaine, MDMA, amphetamines, ketamine, GBH, methamphetamine, mephedrone, and poppers

