## Supplementary material for "Sex differences in health-related quality of life and poverty risk among older people living with HIV in Spain: a cross-sectional study": Sample size calculation

According to the last estimation, 151,387 people living with HIV (PLHIV) in Spain, of which 131,774 know their serological status [1]. Using the latest National hospital survey, we assumed that 53% of PLHIV were 50 years or older [2]. Hence, the total number of PLHIV ≥50 years in Spain (our population of interest) was estimated at 69,840 people.

To make an inference about the health-related quality of life (HRQOL) of PLHIV ≥50 years, we used the standard deviation of ±3.5 (quality of life/general health items) reported for PLHIV ≥50 years in the study conducted by Fuster-Ruiz de Apocada et al. [3]. Assuming a width of the confidence interval of ±0.46 and using a standardised formula for the estimation of means in finite samples with a confidence level of 95%, we obtained a sample size of 222 participants. We used the standard deviation of the average quality of life/general health instead of the mean HRQoL difference between men and women (younger and older participants combined) since literature indicates that there are differences in HRQoL between older and younger people living with HIV [4, 5].

$$n = \frac{N * Z_{\alpha}^{2} * \theta^{2}}{d^{2} * (N-1) + Z_{\alpha}^{2} * \theta^{2}}$$

$$n = \frac{69{,}840 * 1.96^{2} * 12.25}{0.46^{2} * (69{,}840) + 1.96^{2} * 12.25}$$

$$n = 222$$

$\theta$: *standard deviation (general population)*

$N$: *population size(general population)*

$Z_{\alpha}^{2}$: *confidence level*

$d$: *precision level*

Using the most recent Spanish epidemiological surveillance report on HIV and AIDS as a reference [6], we assumed that 84% of HIV infections occurred among men and 16% among women. In order to calculate the number of cases by gender, we used a stratified sample proportionate to the sample size

$$n_h = \frac{N_h}{N} * n$$

$n: stratum\ size\ (general\ population)$

$N: population\ size\ (general\ population)$

$N_h: sample\ size$

The number of questionnaires by gender ($n_h$) is presented in Table S1:

**Table S1. Participants by stratum**

| | Population | Sample |
|---|---|---|
| Men | 58,666 | 186 |
| Women | 11,174 | 36 |
| Total size | 69,840 | 222 |
