## Supplementary material for "Sex differences in health-related quality of life and poverty risk among older people living with HIV in Spain: a cross-sectional study": Psychometric properties of the WHOQoL-HIV BREF questionnaire

In Table S2, we compare the psychometric properties of the WHOQoL-HIV BREF questionnaire from the Spanish validation of the questionnaire [1] with those in our study. We found that Cronbach's alpha (α) coefficients were acceptable (α>0.70) in all domains with the exception of spirituality/personal beliefs (α=0.69). Overall, McDonald's omega (ω) coefficients were more favourable than α measurements. In our study, all domains reported ω values ≥0.81 – except the spirituality/personal beliefs one (ω=0.72). Overall, we found no noticeable differences between the psychometric properties found in the validation study and those in our study.

**Table S2. Psychometric properties of the WHOQoL-HV BREF domains**

| | Cronbach's alpha (α) | | McDonald's omega (ω) | |
|---|---|---|---|---|
| | Validation | Study sample | Validation | Study sample |
| Physical health | 0.73 | 0.81 | 0.79 | 0.81 |
| Psychological health | 0.81 | 0.85 | 0.85 | 0.86 |
| Level of independence | 0.67 | 0.78 | 0.80 | 0.81 |
| Social relationships | 0.75 | 0.81 | 0.81 | 0.82 |
| Environmental health | 0.81 | 0.87 | 0.85 | 0.87 |
| Spirituality/personal beliefs | 0.61 | 0.69 | 0.62 | 0.72 |
